# Application of a Multiplicative Cascade Model to Detect the Early Signs of SARS-CoV-2 Infection Using Heart Rate Data

**DOI:** 10.1101/2022.10.27.22281632

**Authors:** Rachel Heath

## Abstract

**Background:** Wrist-worn devices can keep track of a person’s daily health status, including those likely to become infected with the SARS-CoV-2 virus. Technological solutions using mobile devices are being developed to predict the time course of COVID-19.

**Objective:** In this proof-of-concept study, we use heart rate data to detect the first sign of infection in people who have been diagnosed with COVID-19 and to monitor the time-course of the illness.

**Methods:** The heart-rate data were analysed using a multiplicative cascade driven by a Gaussian process. This provides two parameters, mean and standard deviation, which when combined with similar parameters estimated from control series, provide a Health Index.

**Results:** For 90% of 31 cases, the Health Index tracked COVID-19 infection with the virus and subsequent recovery. The first-sign of COVID-19 was detected on average nine days before symptoms were reported.

**Conclusions:** Early detection of COVID-19 may lead to a reduction in the spread of the virus. The Heath Index’s potential use for the early detection of complications arising from Long COVID would be an important innovation.

## Introduction

Since late 2019, when it first emerged and spread rapidly throughout the world, the SARS-CoV-2 virus has infected millions of people causing illness and sometimes death, while disrupting the social and working lives of many others. Despite the availability of tests for infection with the virus, it would be convenient if an alternate method was possible, one that uses commercially available technology to monitor people passively. Heart rate (HR) is a useful measure that is always available when a wearable device such as an Apple Watch or a Fitbit is being worn. We will review previous research that has used smartwatch HR to detect infection for other diseases and then focus upon a study from the Stanford University Genome Laboratory that has shown promise for predicting the early sign of SARS-CoV-2 virus infection. The example data analyses will use publicly available HR data from the first of these published studies, as for most of the COVID-19 patients the dates on which symptoms first appeared, their diagnosis and recovery are included.

Several medical research organisations have long-term projects investigating the use of mobile and wearable devices for monitoring people’s health. For example, The Scripps Research Institute is conducting a large study using Fitbit and Apple Watch called Digital Engagement and Tracking for Early Control and Treatment (DETECT). Its aim is to use wearable variables to detect and predict the onset of infectious disease. We have shown that HR differences, a proxy for heart rate variability (HRV), can predict the early sign of mental health decline as a steady decrease in the Health Index similar to that used in this paper [1].

Wearable devices can have beneficial effects on health [2]. Short duration HRV measures were collected twice an hour over four weeks from 652 undergraduate students who were also asked to complete rating surveys of their mental health. When the data were analysed separately for day (0800–2400) and night (2400–0800) times using machine learning, correct classification was obtained with 65% success for stress ratings, 72% success for depression and sleep ratings, and 83% success for stomach discomfort, irrespective of the time of day. This result suggests that HR data might be all that is needed to track a person’s wellness over extended periods. HR, as measured by a Fitbit, was related to rated wellness among five of a sample of ten people recruited via the Internet who reported no chronic medical condition [3] using the same technology as was employed in the current study.

### Using Wearable Devices to Monitor COVID-19

About 16% of the US population use a smartwatch. In a population-level study involving Fitbit users from five US states [4], resting HR (RHR) and the incidence of influenza-like illness where these people lived were positively correlated. RHR tends to increase when a person is infected. People infected by the influenza virus show pre-symptomatic signs of an increase in RHR and sleep duration, measures provided by a wearable device linked to a smartphone. HR may be useful for monitoring pre-symptomatic infection in people diagnosed with COVID-19.

Recently, researchers have examined whether HR can be used to detect the early signs of COVID-19 prior to symptoms being reported [5-7], so that people can self-isolate before symptoms appear. Up to 50% of COVID-19 patients can be asymptomatic while infectious[5]. A recent study involved people who owned a smartwatch, such as a Fitbit or an Apple Watch, some of whom were eventually diagnosed with COVID-19 [5]. The data from each participant included their heart rate, steps taken and sleep data, as well as daily symptom reports obtained regularly via a mobile phone app. Of the 4642 participants, 72% wore a Fitbit, 21% wore an Apple Watch and the others wore another type of smartwatch. Of these people, 2.5% reported COVID-19 symptoms and 1% reported other respiratory symptoms.

Using differences in RHR as one of their main dependent variables, the first signs of infection, based on a significant increase in an index derived from RHR differences, occurred on average four days before symptoms were reported and seven days before a formal diagnosis of COVID-19. Detection of COVID-19 infection employed a *cumsum* method that summed the residual RHR measures until a critical value was reached. Using this method, 63% of known COVID-19 infections were detected. A post-hoc analysis of four patient’s HR data showed lingering COVID-19 symptoms long after recovery [5]. So a methodology based on HR might be useful for long-term monitoring of patients diagnosed with Long COVID, a debilitating condition that affects up to 15% of people diagnosed with COVID-19 and can last many months [25-27].

A follow-up investigation [6] developed a real-time COVID-19 infection warning system using HR and step data from a smartwatch. For 80% of the 34 COVID-19 positive patients, pre-symptomatic detection of infection was possible. A novel signal with 80% sensitivity to detect possible infection used deviations from the average RHR recorded overnight. In some cases, these NightSignal deviations occurred from 3 to 10 days before COVID-19 symptoms were reported, thus serving as a suitable warning signal. Of the 18 people who tested positive but had experienced no COVID-19 symptoms, 14 were provided with infection warning signals that had a modal alert time four days before symptoms appeared. However, some of the alarms produced from NightSignal might reflect other causes, different physical conditions, stress and possibly the effects of mental illness episodes [1,6].

We apply a slightly different data analysis method to analyze HR data from Fitbit devices available from [5]. We assume that good health is characterized by a high level of complexity in physiological measures recorded over time, whereas a gradual reduction in complexity signals a decline in health due to infection, as has been shown for mental health relapse [1,8]. Each day’s data for people diagnosed with COVID-19 were analysed using a Gaussian Multiplicative Cascade Model. Technical details of the computations and the statistical method used to detect change over days in parameter estimates are provided in the supplementary files from the OSF Archive for this paper.

### Application of the Multiplicative Cascade Model to Wearable Data

Complexity can be defined in terms of the predictability of a sequence of observations, such as heart rate recorded by a wearable device over a day. Throughout any 24-hr period, HR fluctuates from one moment to the next, the extent of these fluctuations providing information about the complexity of the associated physiological processes. Regular fluctuations indicate a low level of complexity, the least complex example being the regular tick of a clock. As a process becomes more complex its fluctuations become more erratic, a good example being random noise. Most physiological systems exhibit behaviour somewhere between these two extremes. Importantly, research in several domains has found that it is worthwhile monitoring changes in complexity as significant decreases may suggest the onset of pathology [9].

Some medical disorders are characterized by a reduction in the complexity of at least one associated measurement, such as HR in the case of heart dysfunction, EEG amplitude and frequency for epileptic seizures, and mood rating variability for people with unipolar depression [9]. In epileptic seizures, for example, the pre-ictic period prior to the seizure onset is characterized by an increase in slow-wave activity detectable as a reduction in complexity of the corresponding EEG waveform. Using this information, a warning signal can be presented to the person and procedures adopted to avoid a fully expressed epileptic seizure [10]. We propose that a similar diagnostic signal be used to detect changes in a person’s COVID-19 health status using HR data.

### Quantifying Entropy

Entropy is commonly defined as the amount of disorder in a dynamical system. As large values of entropy reflect an increase in the complexity of the underlying dynamic process, quantitative entropy indices serve as useful proxies for complexity in biological systems. *Detrended Fluctuation Analysis* (DFA) has been used to quantify the fractal nature of a time series, such as a series of heart rate measurements. A fractal is a mathematical object that is scale-invariant [11], so that no matter what time scale we choose to measure it, a fractal object always appears the same. We assume that the mathematical form of a HR time series, *HR*(*t*), does not depend on the time scale used to measure it, so it satisfies *HR*(*ct*) =*c*^*H*^*HR*(*t*), where *c* is a scale constant and *H* is the *Hurst Exponent*. The latter is a constant in this *monofractal* case. This self-similarity property has been observed in many physiological time series including HR [12].

A multifractal process involves a generalization of DFA to include processes for which the Hurst exponent is no longer constant over all time scales [13]. Examples from the physical world include atmospheric turbulence and water flowing over a waterfall. While travelling in an aircraft, we experience the effects of atmospheric turbulence on several time scales, moving up and down in your seat, your coffee level bouncing around and the wing ailerons performing a complicated dance to keep the aircraft in level flight.

HR satisfies the mathematical properties of a multifractal process [12,14]. A narrow multifractal spectrum, indicating poor heart function, occurs for people diagnosed with heart failure whereas a wider multifractal spectrum is more likely for people with normal heart function. Long-term HR monitoring of otherwise healthy people may result in useful data related to fluctuations in wellness from one day to the next [14].

### Representing a Multifractal Process as the Output of a Stochastic Multiplicative Cascade

A multifractal process can be represented by a multiplicative cascade that operates on increasingly shorter time scales beginning by dividing the full temporal interval, *T*_*0*_, into two sections at time scale *T*_*1*_, and continuing this division by two at each successive time scale, *T*_*2*_, *T*_*3*_, …, *T*_*k*_. The flow of information from one time scale to the next is determined by the same probability density function, *p*(*x*). Eventually, the hierarchical cascade process produces the HR activity recorded, for example, over a day. The Gaussian Multiplicative Cascade Model (GMCM) is defined when *p*(*x*) is the Gaussian pdf with mean, *μ*, and variance, *σ*^2^,

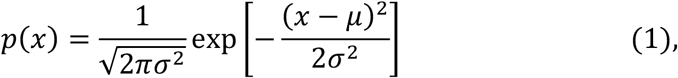

The multifractal spectrum for a binary multiplicative cascade driven by this Gaussian process is given by [17]

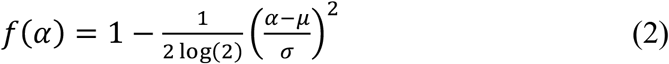

which attains its maximum value of 1 when *α* =*μ*, and its width at *f*(*α*) = 0 is given by 0.2355 *σ*. Estimates of the mean and variance of the Gaussian pdf can be obtained by fitting Eq. 2 to the multifractal spectrum to provide a simple test of the GMCM.

The probability associated with each branch of a multiplicative cascade process can be constructed by applying a logarithmic transform to the product of random variables that are associated with that branch. This results in a random variable equal to the sum of the logarithm of a Gaussian random variable, the latter being represented by a lognormal pdf with parameters *μ* and *σ* [17]. The continuous Shannon entropy, *E*(*μ*, *σ*), for this lognormal pdf is given by

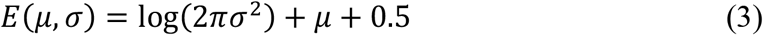

*E*(*μ*, *σ*) is computed for each day’s HR data using the *μ* and *σ* parameter estimates computed from the best-fitting Gaussian multifractal spectrum.

To detect nonlinearity in heart rate data, surrogate comparison series are obtained by computing the power and phase spectra of the original data series, randomly permuting the phase spectrum components, and using the inverse Fourier transform to produce a new time series. This surrogate time series has the same statistical properties as the original time series in terms of its mean, variance, pdf and autocorrelation function, but without any dynamic nonlinearity [18]. For each transformed activity time series, 30 surrogate comparison series were generated using the TISEAN `surrogates` routine [19], available in the `nonlinearTseries` package for R [20]. Evidence for nonlinearity based on the multifractal spectrum was provided by comparing the spectrum produced by the original HR data with a comparison average multifractal spectrum computed using the surrogate data [21].

A Health Index for each day’s HR data can be obtained by computing the standardised Entropy relative to the distribution of Entropy values estimated from the surrogate series. The resulting standardised score, *zEntropy*, measures the multifractal contents of the heart rate series, as follows

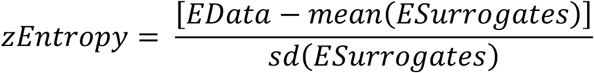

where *EData* is the Entropy estimate from the HR data and *ESurrogates* are the 30 Entropy estimates obtained from the surrogate series [22].

Multifractal analysis can be conducted on HR differences for people diagnosed with COVID-19 using *Multifractal Detrended Fluctuation Analysis* [15,16], as described in more detail in SupplementaryNotes_GMCMDetails.pdf contained in the OSF Archive for this paper. Up to 10,000 HR differences per day were available for analysis to provide accurate estimates of the GMCM predictions.

### Estimating Change Points in a Time Series

A commonly employed statistical technique for detecting change points in a time series estimates future values of the series using a small number of previous values. If the prediction error becomes too large, a change in the parameters governing the time series will have occurred. The method used for this analysis, a semiparametric change detection procedure, can be accessed from commands available in the `sac` R package [23-24]. In this application, the `cumsum.test` command in `sac` using the “epidemic” option was used to detect up to two days on which a change in Health Index is likely, either an increase suggesting improved health or a decrease indicating that things are not going quite so well.

### Application of Fitbit Devices for COVID-19 Monitoring using Heart Rate

Only HR data obtained from a Fitbit smartwatch were used in these analyses due to the much lower density of observations for the Apple Watch. Successive HR values are highly autocorrelated, often up to at least 50 observations into the past. Using successive HR differences generates autocorrelation limited to only one future observation. When a day of HR difference data are analysed using the GMCM, parameter estimates and multifractal spectrum fits are obtained, as shown for a typical case in Figure 1. The close fit of the GMCM to the multifractal spectra is evident by the proximity of the black curve (data) to the black data points and the red curve (average of the surrogate spectra) to the average surrogate spectrum represented by the unfilled red data points. In this example, the estimates of the means for data and surrogate series coincide and the standard deviation of the data series (Gauss) is slightly greater than that for the surrogates (Control). The *zEntropy* estimate is 4.0, suggesting a substantial multifractal component to the heart date differences series recorded from 9,223 heart rate differences obtained on 22 June 2020. This person’s COVID-19 history included the first appearance of symptoms on 14 July, a COVID-19 diagnosis on 17 July and a reported recovery by 6 August.

**Figure 1.**
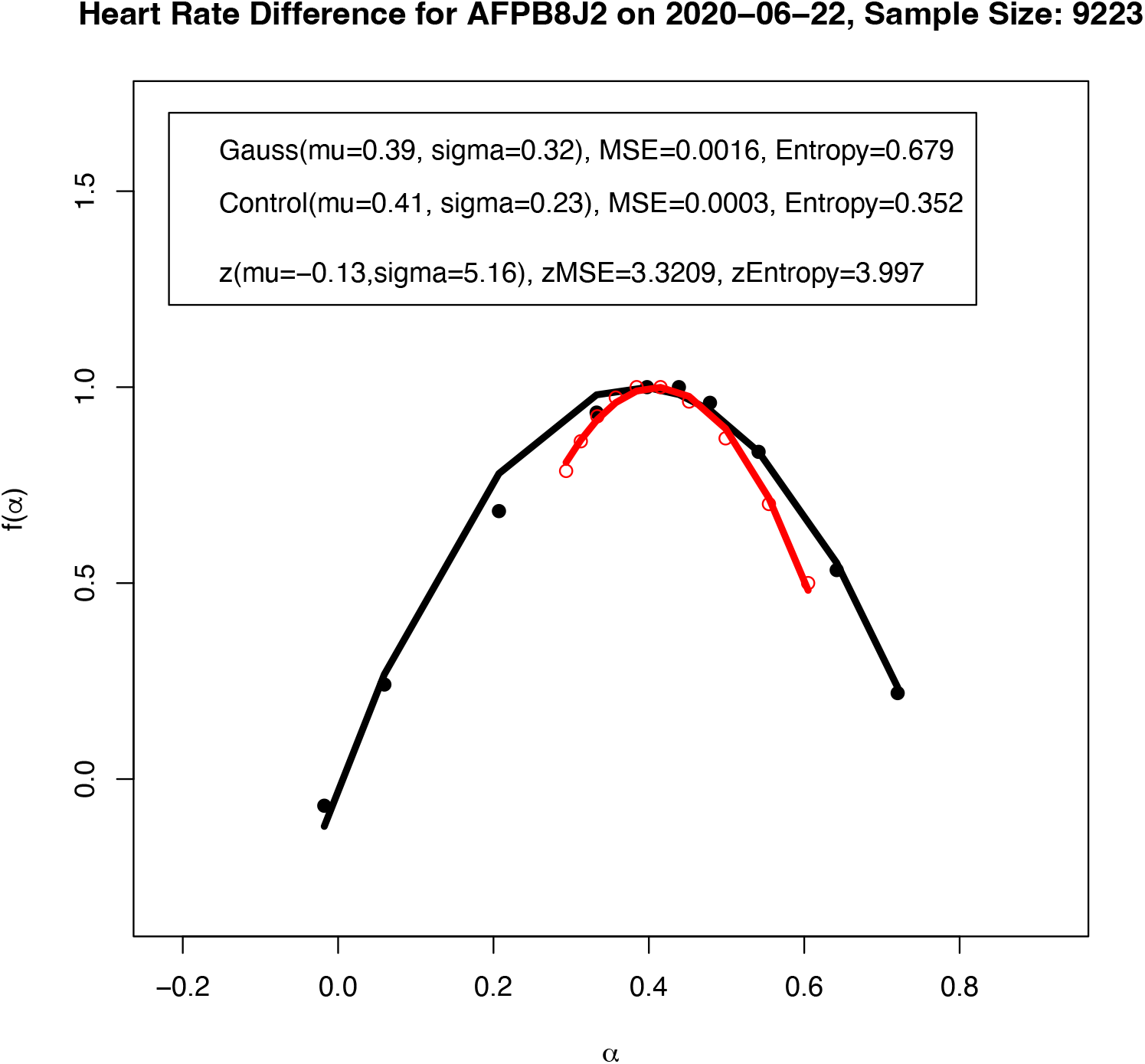
Multifractal spectra for the HR difference data (block dots) and the average of 30 surrogate series derived from the data (red unfilled circles) for COVID-19 case AFPB8J2 [5]. The best-fitting Gaussian Multiplicative Cascade Model is shown by the black curve (data) and the red curve (surrogates).

Figure 2 shows the trends in daily *zEntropy* (top graph) and the sum of a standardized form of *zEntropy* when its values are converted to mean 0 and standard deviation 1 for COVID-19 case AFPB8J2. When the *cumsum* change detection procedure was applied to *zEntropy*, significant changes were detected on 4 July (red dotted line) and 9 August (blue dotted line). A similar *cumsum* analysis of the *Sum*(*zEntropy*) data indicated significant changes on 10 July (dotted orange line) and 24 August (dotted green line). Using *Sum*(*zEntropy*) as a Health Index, we detect the start of declining health on 4 July, a well-established decreasing trend by 10 July, by which time the person had most likely been infected with COVID-19. On 9 August, a low point in the Health Index, a recovery period begins and is well-established by 24 August. A return to this person’s pre-COVID health status does not occur until at least a week later. These dates correspond well with the first sign of infection being detected in the first week of July and recovery being evident about a month later. The COVID-19 time course from the first report of symptoms to recovery is shown by the buff-colored rectangle along the horizontal axis of the *Sum*(*zEntropy*) graph in Figure 2. The first detection of a decline in health status using the *cumsum* change detection method occurred 10 days before COVID-19 diagnosis, an impressive and useful outcome for minimizing the spread of COVID-19 infection.

**Figure 2.**
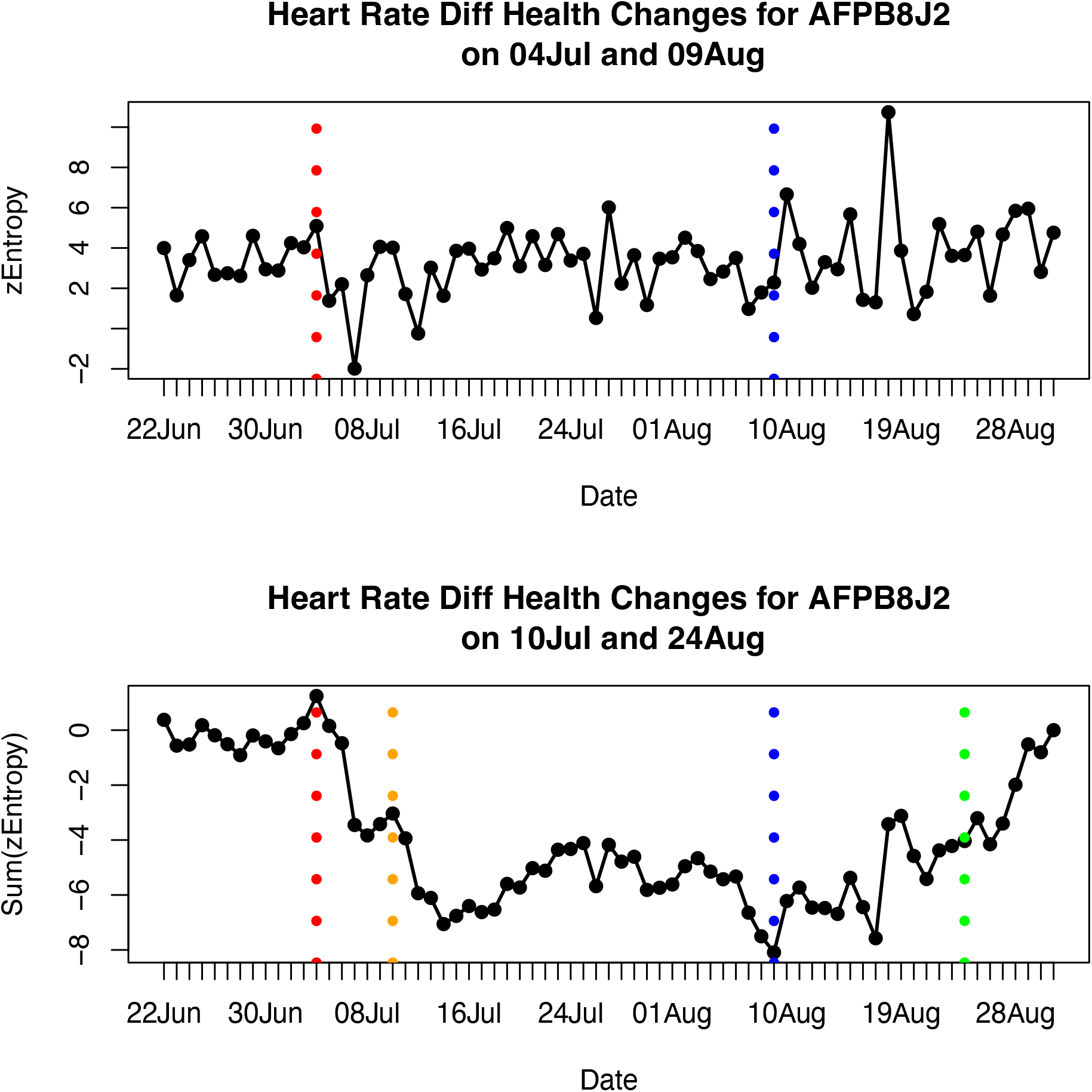
Changes in the *zEntropy* (top graph) and *Sum(zEntropy)* (bottom graph) Health Indices for COVID-19 Case AFPB8J2 [5]. The *cumsum* change detections are represented by the vertical dotted red and blue lines for *zEntropy*, and by the vertical dotted orange and green lines for *Sum*(*zEntropy*). The buff colored rectangle in the bottom graph shows the time-course of COVID-19 from reporting symptoms to recovery.

As *ZEntropy* is better correlated with *sigma* (*r* = 0.69) than *mu* (*r* = −0.10), the prediction of the COVID-19 time course for AFPB8J2 depends more on the rate of spread of activity (*sigma*) in the Gaussian Multiplicative Cascade than its average direction (*mu*). So the illness phase of COVID-19 corresponds to a reduction in the rate of spread of activity in the network, an idea consistent with a reduction in physiological complexity when people are unwell. This effect is demonstrated more clearly when the summed standardized sigma estimates are plotted for this person in Figure 3. There was a rapid decrease in this sum until it reached its minimum when the COVID-19 diagnosis occurred on 15 July. Recovery was evident by the first week of August. Statistical decision thresholds were set at +2 for recovery and at −3 to detect COVID-19 infection, the crossover points indicating possible SARS-Cov-2 infection on 7 July and recovery by 11 August. These dates are close to those obtained using *Sum*(*zEntropy*), as shown in Figure 2.

**Figure 3.**
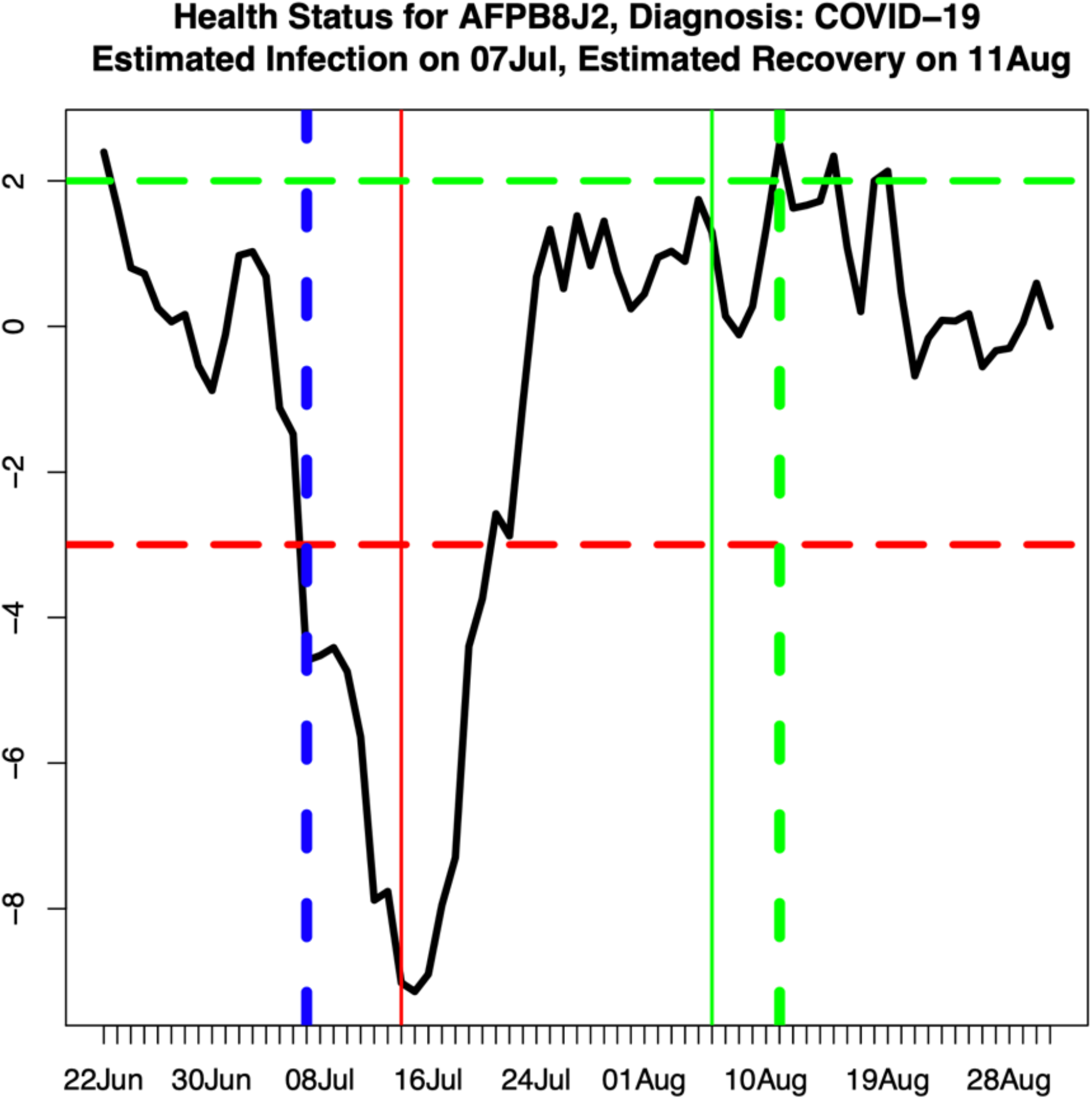
Changes in cumulated standardised *sigma* estimated from the Gaussian Multiplicative Cascade Model applied to heart-rate difference data from COVID-19 case AFPB8J2 [5]. COVID-19 diagnosis occurred on 15 July (vertical solid red line) and recovery occurred on 6 August (vertical solid green line). Positive and negative change detection thresholds were placed at +2 (horizontal dotted green line) and at −3 (horizontal dotted red line), respectively. Using these thresholds, predicted infection occurred on 7 July (vertical dotted blue line) and predicted recovery occurred on 11 August (vertical dotted green line).

Table 1 summarises the analyses for all 31 people who were eventually diagnosed with COVID-19 or at least reported symptoms. *Sum*(*zEntropy*) provided a useful indicator of COVID-19 for all but three of these people, implying a 90% success rate. This represents better performance than the 66% success rate achieved by [5]. The successful predictions have green Case labels in Table 1, and the unsuccessful ones are colored red. Cases labelled L may have experienced an extended period of COVID-19 illness.

**Table 1.**
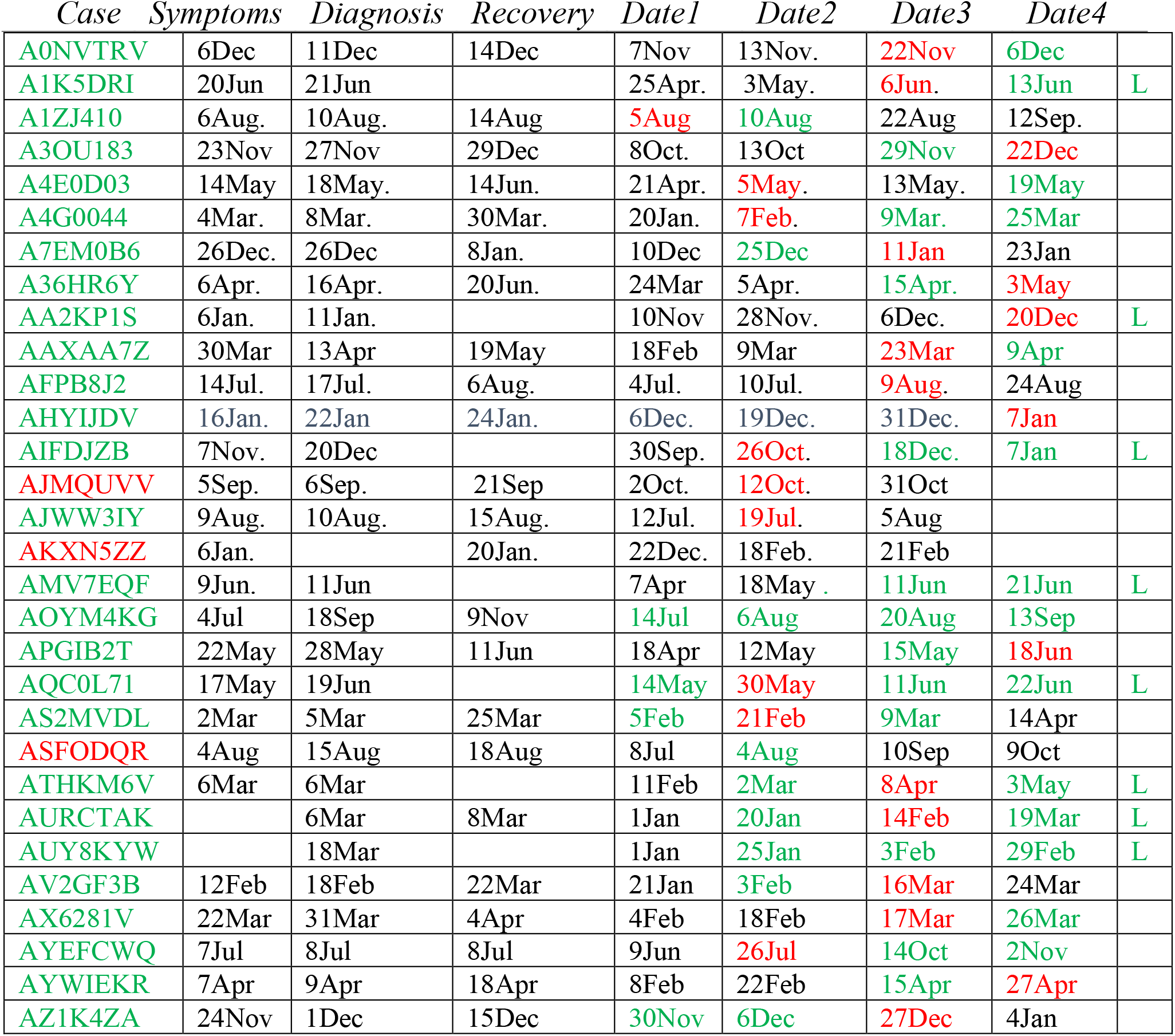
Actual and predicted dates for Symptoms, Diagnosis and Recovery for COVID-19 Cases [5]. Green Case = Prediction was good. Red Case = Prediction was not possible. Red Date1-4 = Minimum of *Sum*(*zEntropy*), Green Date1-4 = Prediction during COVID-19 illness. The L in the right-most column indicates those cases in which the COVID-19 time-course extended until the end of the Fitbit data recording period and possibly beyond.

Table 2 compares the performance of the GMCM model using *zEntropy* and *Sum*(*zEntropy*) to detect change due to COVID-19 infection with performance of the models used by [5]. Both methodologies are based on the same HR data, and comparisons are made with the change detection performance for cases presented in [5, Figure 7e], which shows the relative numbers of detections of change before and after Symptoms are reported for 22 of their cases. Good pre-symptomatic detections are shown in green, post-symptomatic detections are shown in red and some very long pre-symptomatic detections are shown in blue, the latter being considered too long to be effective warnings of future COVID-19 infection. For the 14 cases when a pre-symptomatic alarm was evident, the median delay was nine days before symptom onset. This time compares favorably with the median pre-symptomatic warning of four days provided by the Mishra et al. algorithm [5].

**Table 2.** Comparison of the predictions of the GMCM with [5, Figure 7e]. Cases with good warning before symptoms appeared are shown in green. Post-symptomatic detections are shown in red. The long pre-symptomatic detections are shown in blue.

|  | Mishra et al. 2020 | GMCM analysis |
| --- | --- | --- |
| A0NVTRV | Alarms after Symptoms | Alarm 14 days before Symptoms |
| A1K5DRI | Alarms before and after Symptoms | Alarm 14 days before Symptoms |
| A3OU183 | Alarms before and after Symptoms | Alarm 6 days after Symptoms |
| A4E0D03 | Alarms after Symptoms | Alarm 9 days before Symptoms |
| A4G0044 | Alarms after Symptoms | Alarm 26 days before Symptoms |
| A7EM0B6 | Alarms after Symptoms | Alarm 1 day before Symptoms |
| AA2KP1S | Alarms before and after Symptoms | Alarm 17 days before Symptoms |
| AAXAA7Z | Alarms before and after Symptoms | Alarm 7 days before Symptoms |
| AHYIJDV | Alarms after Symptoms | Alarm 9 days before Symptoms |
| AIFDJZB | Alarms before and after Symptoms | Alarm 11 days before Symptoms |
| AJWW3IY | Alarms before and after Symptoms | Alarm 4 days before Symptoms |
| AKXN5ZZ | Alarms after Symptoms | No alarm before Symptoms |
| AMV7EQF | Alarms before and after Symptoms | Alarm 2 days after Symptoms |
| AOYM4KG | Alarms before and after Symptoms | Alarm 9 days before Symptoms |
| APGIB2T | Alarms after Symptoms | Alarm 10 days before Symptoms |
| AQC0L71 | Alarms after Symptoms | Alarm 18 days before Symptoms |
| AS2MVDL | Alarms after Symptoms | Alarm 10 days before Symptoms |
| ASFODQR | Alarms after Symptoms | No alarm before Symptoms |
| AV2GF3B | Alarms after Symptoms | Alarm 9 days before Symptoms |
| AX6281V | Alarms before and after Symptoms | Alarm 5 days before Symptoms |
| AYEFCWQ | Alarms before and after Symptoms | Alarm 29 days before Symptoms |
| AYWIEKR | Alarms after Symptoms | Alarm 8 days after Symptoms |

A comparison of the respective technologies can be obtained by forming the cross-classification of COVID-19 Symptom prediction success and failure for the two methods as shown in Table 3. The GMCM predicted a decline in health prior to Symptoms being reported for 18 of out of 22 cases (82%) compared with 10 out of 22 cases with the Mishra et al. [5] technology (45%). The better performance of the GMCM model is indicated by a significant Chi-squared Test using the frequencies in Table 3, X^2^(1 *df*) =4.81, *P* =.028. However, for four of the cases that were correctly predicted by the GMCM, predictions occurred between 17 and 29 days before symptoms were reported. If these predictions are reclassified as failures for the GMCM, there would be no significant difference in the COVID-19 prediction performance of the GMCM and the Mishra et al. techniques, X^2^(1 *df*) =2.35, *P* =.125.

**Table 3.** Comparison of the performance of the GMCM with the technology used by Mishra et al. [5]

| Prediction | GMCM | Mishra et al.[5] |
| --- | --- | --- |
| Before Symptoms | 18 | 10 |
| After Symptoms | 4 | 12 |

## Conclusions

An analysis of the Mishra et al. [5] HR data has shown that the GMCM plus its *cumsum* change detection procedure can detect evidence of COVID-19 infection before symptoms are reported. The accuracy of GMCM is similar to the data analysis method of Mishra et al. that uses a transformation of HRV data and a similar type of sequential decision making scheme. For those cases providing adequate data, the GMCM provided an average pre-symptomatic warning about five days earlier than did the Mishra et al. data analysis method. These extra five days are crucial when potential COVID-19 sufferers might otherwise self-isolate and not be moving around spreading the virus among the community.

It is better that a person’s HR data from a wearable device be analysed in real time 24/7 using a special mobile device application, so that they can be advised to take a COVID-19 test well before symptoms appear. A modification of the research tool, *PhDApp*, could perform this task [6]. A significant reduction in the *Sum*(*zEntropy*) Health Index could result from causes other than Sars-CoV-2 infection, such as declining mental health and imminent relapse in long-term conditions such as mood disorder, [1,8]. There are also the normal ups and downs anyone can experience, such as an improvement in health with good diet and exercise and a temporary decline when one is not adjusting too well to environmental stresses, or when a person undergoes an invasive medical procedure [3]. The GMCM and associated change detection technology may find useful application in monitoring the health fluctuations in people diagnosed with Long COVID, a persistent form of the illness that can affect numerous body functions, including cognition [25-27].

## Data Availability

All data produced are available online at
https://osf.io/mwnx3/

https://storage.googleapis.com/gbsc-gcp-project-ipop_public/COVID-19/COVID-19-Wearables.zip

## Acknowledgments

We thank the members of the Stanford University Genomics Laboratory for allowing us to use their publicly available data sets. We adjusted the year in these data files to be 2020.

## Multimedia Appendix

Data URL: https://storage.googleapis.com/gbsc-gcp-project-ipop_public/COVID-19/COVID-19-Wearables.zip

The supplementary files are contained in the OSF archive directory https://osf.io/mwnx3/ The GMCM is described in more detail in SupplementaryNotes_GMCMDetails.pdf The GMCM parameter estimates are contained in Results_CovidPredictionUsingHeartRate.zip The Figures for all participants are contained in “SupplementaryFile_Extra Figures.pdf”

## Abbreviations

DFA: detrended fluctuation analysis
EEG: electroencephalogram
GMCM: gaussian multiplicative cascade model
HR: heart rate
HRV: heart rate variability
pdf: probability density function
RHR: resting heart rate

